# Prevalence and correlates of urogenital schistosomiasis in school going children in Maramba compound of Livingstone District, Zambia

**DOI:** 10.1101/2022.04.08.22273629

**Authors:** Shike Kapanga, John Amos Mulemena, Kingsley Kamvuma, Christopher Newton Phiri, Warren Chanda

## Abstract

**Background:** Schistosomiasis is an acute and chronic parasitic disease that is caused by trematode worms (blood flukes) of the genus Schistosoma. *Schistosoma haematobium* (*S. haematobium*) is known to cause urogenital schistosomiasis. The disease is the second most common socio-economically devastating tropical parasitic disease after malaria in Africa. In Zambia, it affects over a million school going children, mostly in rural communities due to unsafe water and inadequate sanitation facilities. This study aimed to determine the presence of *S. haematobium* in urine specimens of school going children in Maramba compound of Livingstone and establish factors associated with the acquisition and spread of the parasite.

**Methods:** A structured questionnaire was administered on all children with signed consent from their guardians/parents and afterward spot urine specimens were collected in sterile containers for macroscopically/microscopically examination by an independent laboratory technologist.

**Results:** A total of 173 school going children participated in the study. Parasitic eggs were detected in 6 specimens providing a prevalence of 3.47% (p<0.01) and this had a strong association with presence of microscopic red blood cells (p<0.01), dysuria (p=0.026), washing in a stream (p=0.01), and the perception on bilharzia acquisition (p<0.01).

**Conclusion:** The prevalence of urogenital schistosomiasis among school going children in Maramba compound was 3.47%, and the correlates of the infection included washing in a stream, older age and poor knowledge on schistosomiasis. Participants that had schistosomiasis often presented with hematuria and lacked knowledge on disease acquisition, health effects and preventive measures. This calls for more robust sensitization of school going children and periodic screening to curb the disease.

## Introduction

Schistosomiasis is an acute and chronic parasitic disease that is caused by trematode worms (blood flukes) of the genus Schistosoma. This genus causes 2 major forms of schistosomiasis (intestinal and urogenital) with 5 main species of blood flukes such as *Schistosoma mansoni, Schistosoma japonicum, Schistosoma mekongi, Schistosoma guineensis, and Schistosoma haematobium; and* the three most common are *S. haematobium, S. japonicum, and S. mansoni* (1, 2). *Schistosoma haematobium* is known to cause urogenital schistosomiasis. The disease is the second most common socio-economically devastating tropical parasitic disease after malaria in Africa (3). Schistosomiasis infects over 230 million people annually and causes a worldwide burden of over 3 million disability-adjusted life years (4-6). The World Health Organization (WHO) recognizes schistosomiasis as a Neglected Tropical Disease (NTD) with endemicity in 78 tropical and sub-tropical countries (7, 8), thus making it a disease of public health concern.

Studies have indicated that exposure to infested water by people during routine agricultural, domestic, occupational, and recreational activities, is the main channel of disease transmission (9, 10). The life cycle of schistosome occurs in snails and mammals. In the snail, the process starts with the development of miracidia into a sporocyst which later grow and multiply into cercariae, the developmental stages that further grow and mature into full grown parasites that mate and produce eggs in mammalian hosts (2). Through urine or feces, eggs are released into the environment from mammalian hosts and hatch into miracidia that subsequently infect snails in fresh water (11).

Children are at increased risk of contracting the disease. Certain recreational activities such as swimming or fishing in infested water and lack of hygiene put school-aged children vulnerable to infection (3). Male and female Schistosoma worms have the ability to avoid the host immunity and can localize in the pelvic venous plexus for decades. Eggs deposited in the bladder wall are resistant to T-helper-1(Th1) cell elimination through formation of fibrous capsule and progressive bladder disease which leads to complications such as obstructive uropathy and predisposition to squamous cell carcinoma (3).

Globally, the prevalence of schistosomiasis has been associated with poor sanitary conditions and consumption of untreated water. In Africa, several studies indicate the increased prevalence of the disease in school-going children. For instance, 57.6% prevalence was reported in 12 villages of Niakhar region of Fatick, Senegal among school-going children aged 7-15 years (12), a mean prevalence of *S. haematobium* was 30.7% in Zanzibar (13), and the prevalence of 52.0% was recorded in age group 11 – 20 years in a rice-farming community in Adim Village in Biase area of Cross River State, Nigeria (14). These studies suggested that the rural communities in African countries are mostly affected.

In Zambia, rapid changes in environmental factors and social economic factors are linked to infections due to *S. haematobium*. The infection affects more than 4 million school-aged children, mostly in rural communities due to unsafe water and inadequate sanitation facilities (15). A meta-analysis study revealed a 32.2% prevalence estimate of *S. haematobium* across studies for the entire period among school-aged children (15). Despite the implementation of massive drug administration (MDA) by the Ministry of Health to curb the disease, sporadic cases are being reported by various hospitals in Zambia. Thus, periodic monitoring of the program is important to understand the situational status. Maramba compound(Figure 1) in the town of Livingstone, is a peri-urban area attached to the stream with a total population of 9590, of which 2747 (29%) were children aged 10-19 years.(16) The stream is used for various human activities such as gardening swimming, fishing, washing etc. and we hypothesized that it might be infested with Schistosoma vectors. Therefore, this study aimed to determine the presence of *S. haematobium* in urine specimens of school going children in Maramba compound of Livingstone and establish factors associated with the acquisition and spread of the parasite.

**Figure 1:**
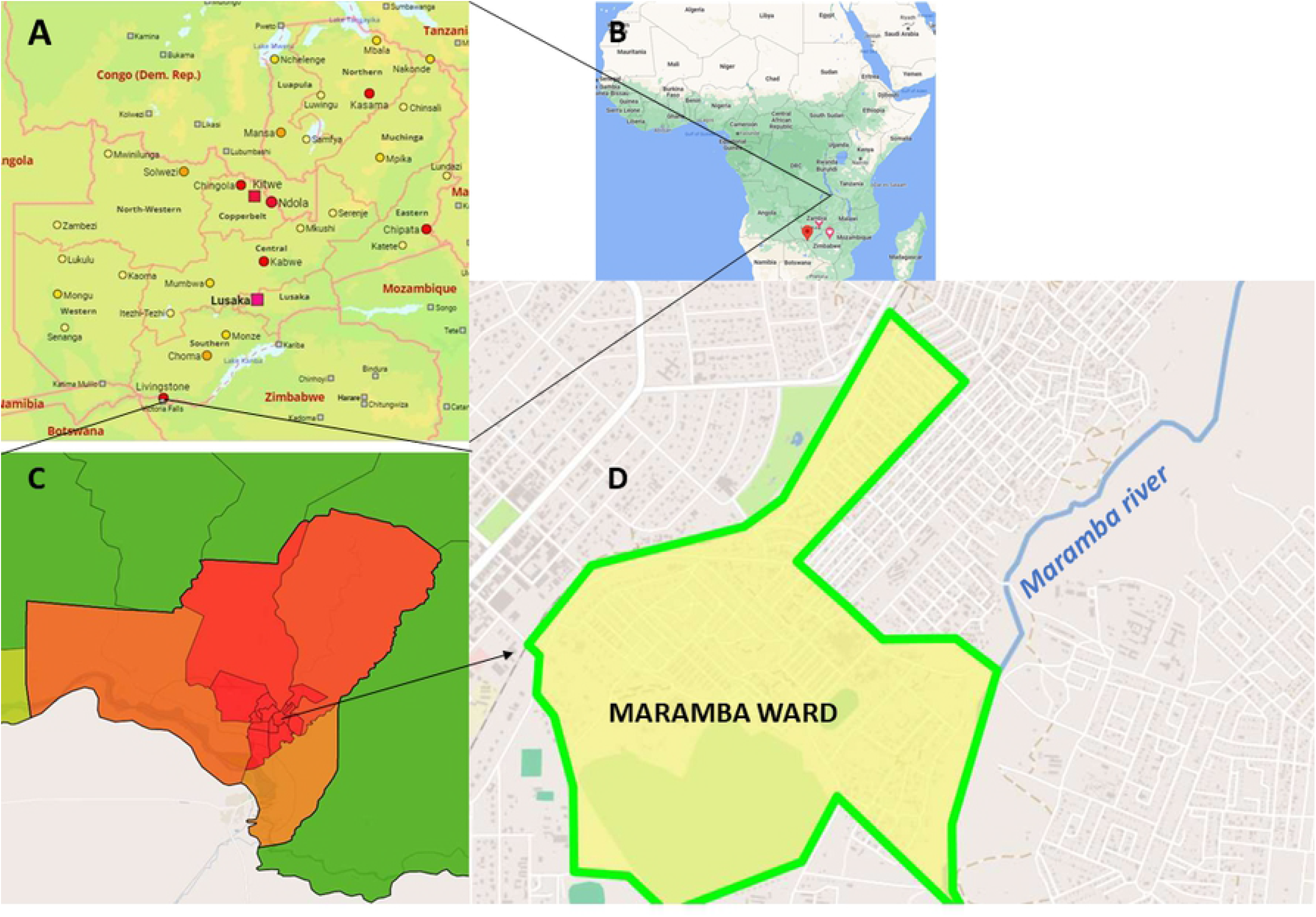
The geographical location of Maramba compound. The map of Zambia (A) zoomed from the map of Africa (B), map of Livingstone city (C), and the study site-Maramba compound with Maramba river (D). courtesy of Thomas Brinkhoff (16)

## Methods and Materials

A cross-sectional study was conducted on school going children at Maramba Primary School in Maramba compound of Livingstone city after obtaining ethical clearance (SMHS-MU2-2020-21) from Mulungushi University School of Medicine and Health Sciences Research Ethics committee and permission from Livingstone District Health office. The School registers were used to randomly select participant using a systematic sampling method. From the register, every fourth name was selected for the study and all pupils (6-16years old) that visited the out-patient department at Maramba Clinic were also randomly selected for the study.

A structured questionnaire was administered on all children with signed consent from their guardians/parents and afterward spot urine specimens were collected in sterile containers for macroscopically/microscopically examination by an independent laboratory technologist. However, school going children whose guardians/parents refused to sign the consent form, or had taken medication for schistosomiasis 2 weeks prior to sample collection were excluded from the study.

The outcome study variable was the detection of *S. haematobium* eggs in urine specimen while the independent variables included age, gender, water source, sanitation levels, proximity to stream, swimming in contaminated water, previous UTI diagnosis, UTI medication history, headache, fever, myalgia, rash, dysuria, and microscopic red blood cells in urine.

Data was entered, cleaned and coded in Microsoft Excel 2019 then exported and analyzed using STATA version 15. Descriptive statistics using tables, frequencies, means and medians were utilized to describe the data. A chi-square test was used for categorical variables and logistic regression was used to analyze the logarithmic transformation of the odds of the associated variables. A p-value of ≤5% was considered statistically significant.

## Results

### Demographic and clinical characteristics

A total of 173 school going children participated in the study. Out of 173, 57.23% (99) were females and 42.77% (74) were males with the age ranging from 10-16 years (mean age: 12). Among the participants, 98.27%, (170) had no history of urinary tract infection (UTI), 97.69% (169) had no history of UTI medication, 72.83% (126) took undisinfected drinking water, 91.91% (159) had piped water, 63.01% (109) used flushable toilets, and 82.66% (143) had no history of swimming in the stream as presented in Table 1.

**Table 1:** Frequencies of demographic and clinical characteristics of the study participants.

| Variable | Mean (IQR) | Frequency | Percent (%) |
| --- | --- | --- | --- |
| **Age child** | 12.9(10,16) |  |  |
| **Gender child** |  |  |  |
| Male |  | 74 | 42.77 |
| Female |  | 99 | 57.23 |
| **Previous UTI diagnosis** |  |  |  |
| Yes |  | 3 | 1.73 |
| No |  | 170 | 98.27 |
| **UTI medication history** |  |  |  |
| Yes |  | 3 | 1.73 |
| No |  | 170 | 98.27 |
| **Toilet Used** |  |  |  |
| Latrine |  | 64 | 36.99 |
| Flushable toilet |  | 109 | 63.01 |
| No Toilet |  | 0 | 0 |
| **Source of Water** |  |  |  |
| Stream |  | 2 | 1.16 |
| Tapwater |  | 159 | 91.91 |
| Borehole |  | 12 | 6.94 |
| Well |  | 0 | 0 |
| **Disinfecting Drinking Water** |  |  |  |
| Yes |  | 47 | 27.17 |
| No |  | 126 | 72.83 |
| **Living near a stream** |  |  |  |
| Yes |  | 28 | 16.18 |
| No |  | 145 | 83.82 |
| **Washing in a Stream** |  |  |  |
| Yes |  | 12 | 6.94 |
| No |  | 161 | 93.06 |
| **Swimming in a Stream** |  |  |  |
| Yes |  | 30 | 17.34 |
| No |  | 143 | 82.66 |
| **Perception on Bilharzia acquisition** |  |  |  |
| eating unwashed foods |  | 3 | 1.73 |
| walking barefoot in contaminated soil |  | 8 | 4.62 |
| contact with contaminated water |  | 123 | 71.1 |
| mosquito bite |  | 7 | 4.62 |
| no response |  | 26 | 15.03 |
| Other (urinating in dirty water) |  | 6 | 3.47 |
| **Perception on Bilharzia Health Effects** |  |  |  |
| blood loss |  | 73 | 42.2 |
| anemia & malnutrition |  | 22 | 12.72 |
| disturbs children's growth |  | 20 | 11.56 |
| disturbs learning well |  | 13 | 7.51 |
| causes chronic severe disease |  | 30 | 17.34 |
| no response |  | 15 | 8.67 |
| **Perception on Infection Prevention** |  |  |  |
| Defecating/urinating in latrines |  | 22 | 12.72 |
| Avoid bathing in contaminated water |  | 72 | 41.62 |
| wash hands before eating |  | 42 | 24.28 |
| Other |  | 35 | 20.23 |
| no response |  | 2 | 1.16 |
| **Past Bilharzia diagnosis** |  |  |  |
| Yes |  | 16 | 9.25 |
| No |  | 157 | 90.75 |
| **Headache** |  |  |  |
| Yes |  | 42 | 24.28 |
| No |  | 131 | 75.72 |
| **Fever** |  |  |  |
| Yes |  | 31 | 17.92 |
| No |  | 142 | 82.08 |
| **Body aches** |  |  |  |
| Yes |  | 27 | 15.61 |
| No |  | 146 | 84.39 |
| **Rash** |  |  |  |
| Yes |  | 7 | 4.05 |
| No |  | 166 | 95.95 |
| **Dysuria** |  |  |  |
| Yes |  | 15 | 8.67 |
| No |  | 158 | 91.33 |
| **History of hematuria** |  |  |  |
| Yes |  | 19 | 10.98 |
| No |  | 154 | 89.02 |
| **Anemic** |  |  |  |
| Yes |  | 7 | 4.05 |
| No |  | 166 | 95.95 |
| **Antibiotics** |  |  |  |
| Yes |  | 1 | 0.58 |
| No |  | 172 | 99.42 |
| **Deworming medication** |  |  |  |
| Yes |  | 26 | 15.03 |
| No |  | 147 | 84.97 |
| **Microscopic red blood cells** |  |  |  |
| Yes |  | 6 | 3.47 |
| No |  | 167 | 96.53 |
| **Schistosoma cyst in urine** |  |  |  |
| Yes |  | 6 | 3.47 |
| No |  | 167 | 96.53 |

Studies have revealed that trapped parasitic eggs induce a distinct immune-mediated granulomatous response that causes pathological effects ranging from anemia, growth stunting, impaired cognition, and decreased physical fitness, to organ-specific effects such as severe hepatosplenism, periportal fibrosis with portal hypertension, and urogenital inflammation and scarring (17). Few weeks after infection, some people may present with fever, an itchy and raised rash, muscle and joint pain, and general body malaise (18, 19). In our study, the majority of participants were asymptomatic; afebrile, no headache, no dysuria and body aches, no skin rash and were not anemic (Table 1).

Moreover, we assessed the children’s perception towards bilharzia acquisition, health effects and infection prevention. Despite bilharzia lessons being taught in primary education science subjects, 71.1% (127) indicated that bilharzia could be acquired through contact with contaminated water while 15.03% (26) declined to respond (Table 1). On health effects associated with bilharzia, blood loss stood out the most with 42.2% (73) whereas avoiding bathing in contaminated water (41.62%, 72) and handwashing before eating (24.28%, 42) were identified as infection preventive measures (Table 1).

Laboratory findings revealed presence of red blood cells and Schistosoma cysts in 3.47% (6) of the urine specimens analyzed (Table 1).

### Prevalence of *Schistosoma haematobium* infection and related factors

The presence of Schistosoma cysts in urine denoted urogenital schistosomiasis infection. The cysts were detected in 6 specimens providing a prevalence of 3.47% (p<0.01) and this had a strong association with presence of microscopic red blood cells (p<0.01), dysuria (p=0.026), washing in a stream (p=0.01), and the perception on bilharzia acquisition (p<0.01) as presented in Table 2.

**Table 2:** Prevalence and correlates of *Schistosoma haematobium* infection.

| Variable | Number | S. haematobium |  | P-value |
| --- | --- | --- | --- | --- |
|  |  | Yes<br>6(3.47%) | No<br>167(96.53%) |  |
| Age child Mean (IQR) |  | 15(13,16) | 13(10,16) | 0.4 |
| Washing in a Stream | 173 |  |  | 0.01 |
| Yes |  | 2(33.33) | 10(5.99) |  |
| No |  | 4(66.67) | 157(94.01) |  |
| **Perception on Bilharzia Acquisition** | 173 |  |  | **<0.01** |
| eating unwashed foods |  | 0 | 3(1.80) |  |
| walking barefoot in contaminated soil |  | 0 | 8(4.79) |  |
| contact with contaminated water |  | 0 | 123(73.65) |  |
| mosquito bite |  | 1(16.67) | 6(3.59) |  |
| no response |  | 2(33.33) | 24(15.03) |  |
| Other (urinating in dirty water) |  | 3(50) | 3(3.47) |  |
| **Perception on Bilharzia Health Effects** | 173 |  |  | **0.026** |
| blood loss |  | 1(16.67) | 72(43.11) |  |
| anemia & malnutrition |  | 0 | 22(13.17) |  |
| disturbs children's growth |  | 0 | 20(11.98) |  |
| disturbs learning well |  | 2(33.33) | 11(6.59) |  |
| causes chronic severe disease |  | 1(16.67) | 29(17.37) |  |
| no response |  | 2(33.33) | 13(7.78) |  |
| **Dysuria** | 173 |  |  | **0.029** |
| Yes |  | 2(33.33) | 13(7.78) |  |
| No |  | 4(66.67) | 154(92.22) |  |
| **Hematuria** | 173 |  |  | **<0.01** |
| Yes |  | 6(100) | 13(7.78) |  |
| No |  | 0 | 154(92.22) |  |
| **Microscopic red blood cells** | 173 |  |  | **<0.01** |
| Yes |  | 6(100) | 0 |  |
| No |  | 0 | 167(100) |  |
| **Schistosoma cyst in urine** | 173 |  |  | **<0.01** |
| Yes |  | 6(100) | 0 |  |
| No |  | 0 | 167(100) |  |

Among the 6 participants that had schistosomiasis 33.33% (2) washed their clothes in a stream (p=0.01) while 66.67% (4) did not. On the acquisition of the disease, 50% (3) mentioned obtaining it via urinating in dirty water, 33.33% (2) declined to respond while 16.67% (1) indicated as being through mosquito bites (Table 2). However, participants without the disease thought that coming into contact with contaminated water was a way of contracting bilharzia (73.65%, 123; p<0.01).

Understanding the health effects that bilharzia impact on affected individuals may help in identifying it sooner and cause them to seek medical attention. In our study, participants who were negative for the disease indicated that blood loss (43.11%, 72) is a health effect of bilharzia whereas amongst the 6 participants detected with bilharzia; health effects chosen were blood loss (16.67%, 1), disturbs learning (33.33%, 2), causes chronic severe disease (16.67%, 1), but 33.33% (2) declined to respond (p=0.026, Table 2).

### Univariable and Multivariable analysis of factors associated with schistosomiasis

During univariable analysis it was found that an increase in age was associated with an increased chance of having schistosomiasis (OR 1.22;95%CI: 1.41-5.84, p= 0.004) as shown in Table 3. Although statistically insignificant, participants who were male had an increased chance of acquiring schistosomiasis as compared to those that where female (OR 2.77; 95%CI: 0.49-15.56, p=0.247). It was also discovered that people that never washed their clothes in the stream had reduced odds of having schistosomiasis as compared to those that washed clothes in the stream (OR 0.127; 95%:CI 0.021-0.781, p=0.026).

**Table 3:**
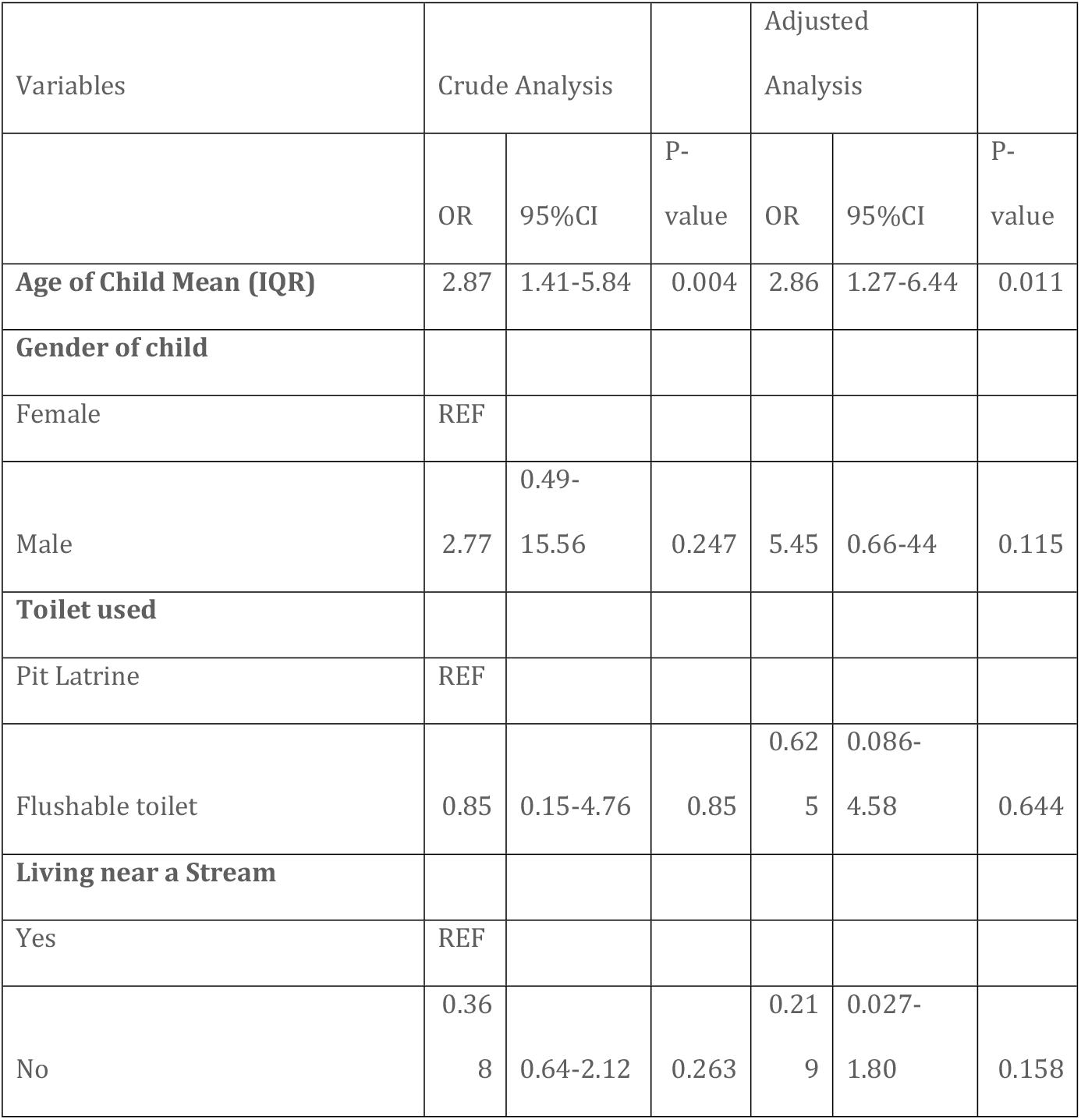
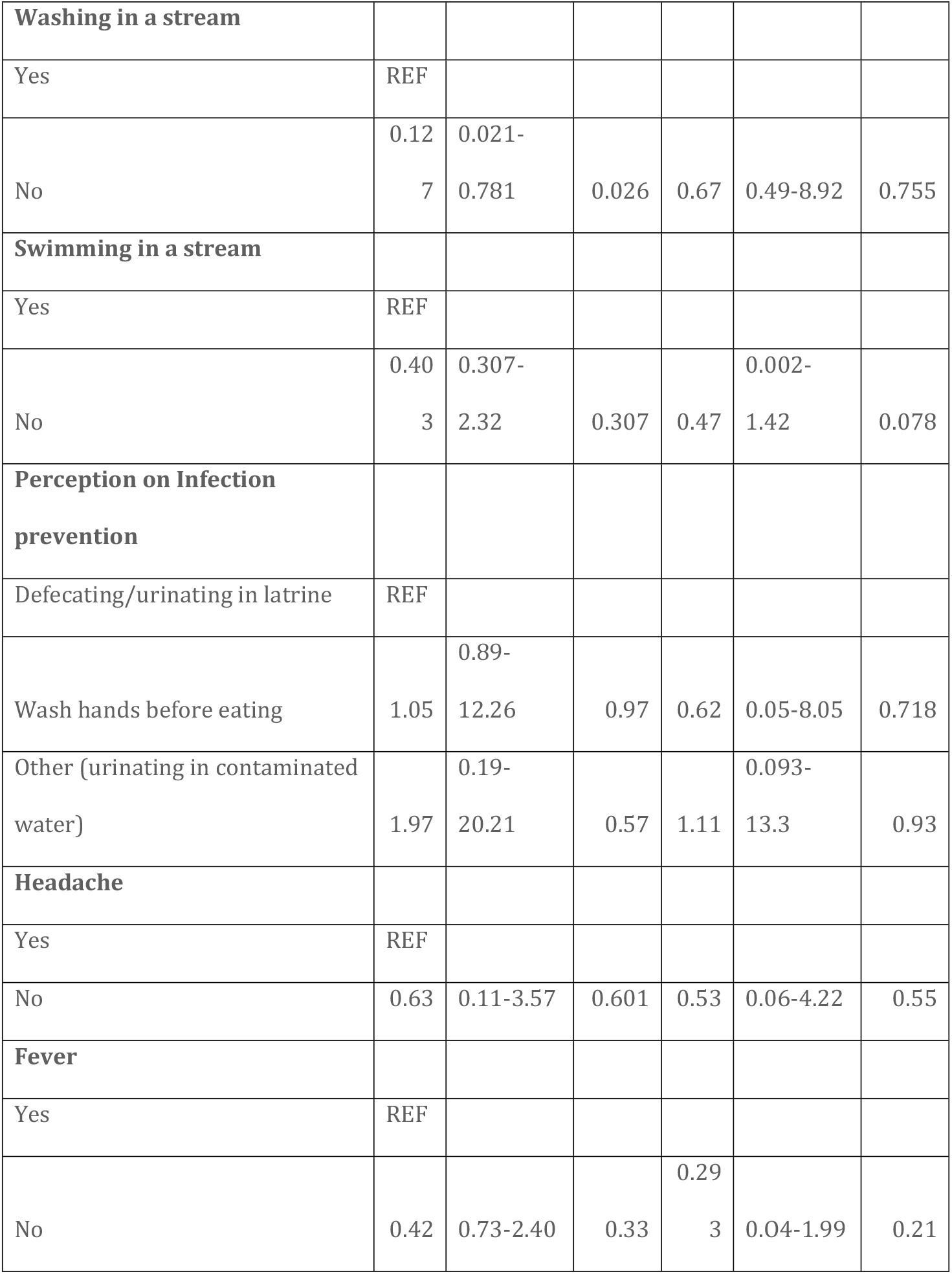
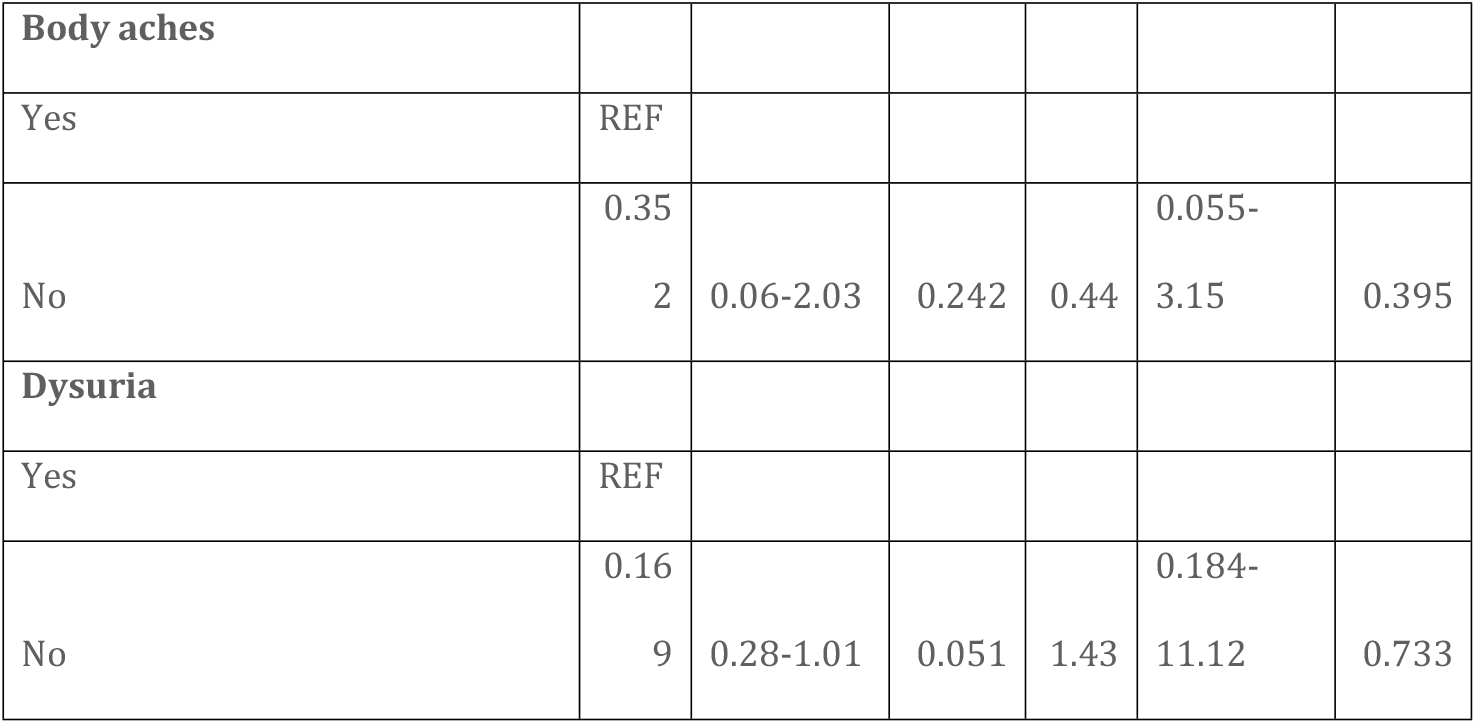
Univariable and multivariable analysis of correlates of schistosomiasis.

During multivariable analysis only age was found to be statistically significant (p=0.011). It was observed that an increase in age was associated with an increased odds of acquiring schistosomiasis (OR 2.86; 95%CI: 1.27-6.44; p= 0.011), this was supported by a linear regression analysis (R2: 0.063, p=0.001).

## Discussion

Urogenital schistosomiasis is a neglected tropical disease caused by *Schistosoma haematobium* that resides in the vasculature surrounding the urogenital system, and is associated with perturbations in the gut microbiota (20). The disease is prevalent in several parts of Africa especially in areas with large water bodies. Exposure to infested water by people during routine agricultural, domestic, occupational, and recreational activities, is the main channel of disease transmission (9, 10). The infection affects more than 4 million school-aged children, mostly in rural communities due to unsafe water and inadequate sanitation facilities (15). Mass treatment of school children has been a regular exercise often undertaken by governments and stakeholders to decrease the disease burden and reduce transmission in selected communities.

Urogenital schistosomiasis can have unacknowledged devastating impact on the urinary tract that could have implication for progressive renal damage which, if not detected and treated, could lead to end stage renal failure and death (21). Looking at the recreational swimming in school going children as a potent medium for the spread of disease that is usually asymptomatic (3), periodic screening are warranted to protect the lives of children and adolescents in communities. Therefore, this study was conducted to assess the prevalence of urogenital schistosomiasis and to understand associated factors in Maramba compound of Livingstone city. The study targeted school going children with age ranging from 10-16 years old. The Compound has a stream that children swim from, and is used as a source of water for gardening and washing of clothes. We therefore, hypothesized that school going children may be unregulated to utilize the stream for various recreational activities and contract the parasites. Our findings showed a low prevalence of 3.47% which was consistent with similar studies done in Egypt (22) and Ghana (23). However, our prevalence rate was lower than the 5.3% and 13.9% prevalence rates discovered by Halwindi *et al*.(24) in Mazabuka and Siavonga districts of Zambia, respectively. The variations could be attributed to differences in the seasons of data collection, geographical location, and the sample sizes as they considered five catchment areas while our study only considered one area with low sample size (comprising 173 participants). A review study(25) that considered related studies in the central and southern regions of Africa discovered a prevalence of 73.3%, 47.0% and 33.2% schistosomiasis among school aged children in the Eastern Cape province of South Africa, Mozambique and Sengerema district of North-West Tanzania, respectively. Despite several African countries employing the massive chemotherapy with praziquantel to contain and control the infection, a plethora of literature revealed the presence of the disease in several communities. The questions that beg answers would be on the effectiveness of the massive treatment program. Should implementers change the strategies of massive drug administration (MDA) or how potent are these drugs? Is the pathogen developing resistance or should MDA programs be simultaneously be implemented with vector control programs in high-risk communities? These questions can only be answered when implementational research and antimicrobial resistance studies are conducted putting into consideration that praziquantel rarely eliminates infection but reduces morbidity and might impact on transmission (26, 27).

A study by Gbalégba et al. (28) found males to be nearly two times more likely to be infected with *S. haematobium* than females (adjusted OR, (aOR) 1.75, 95% CI: 1.11–2.77), and children at primary school were nearly two times more likely to be infected than those who did not go to school (aOR 1.79, 95% CI: 1.07–2.99). Our study involved children at primary school and the male gender was not strongly associated with contracting *S. haematobium* after confounding factors were excluded.

In establishing correlates of schistosome infections, variables such as human contact with water and socio-demographics create a high risk of exposure (29). Our study found that not living near a stream, not washing in a stream and avoiding swimming in a stream reduced chances of putting children at risk of contracting schistosome infection by 22%, 67% and 47%, respectively. This was in line with a study done in Mauritania which found swimming/bathing as the main activity, followed by washing clothes and dishes (28). The variability of ambient temperature and humidity in sub-Saharan Africa forces children to have swimming activities and general interaction with water. Therefore, mollusk control programs might prevent disease spread since contact with water may not be avoided by children.

Furthermore, we discovered that an increase in age was associated with an increased odds of acquiring *S. haematobium* (aOR 2.86; 95% CI: 1.27-6.44, *p*=0.011) and this was similarly seen in a study in Yemen where the prevalence of *S. haematobium* infection was found significantly higher in children aged over 10 years and lower in younger ones (30). This observation could probably be because this age group is often involved in water-contact activities such as swimming, washing, watering or fishing.

Despites clinical manifestations such as headache, fever, skin rash and dysuria having an association with schistosomiasis, our study found a reduced connection with the infection after a multivariate analysis. However, hematuria had a strong connection with the infection because infected participants had it. This was in part similar to a study done elsewhere (31) which suggested that hematuria presence could act as a predictor of bilharzia especially in endemic regions.

Understanding the health behavior of the target population is key for curbing schistosomiasis. In a rural area of KwaZulu-Natal, a study showed that basic knowledge about the risk of schistosomiasis among the participants was high, but the cause and prevention of the disease were less well understood (32). Elsewhere in Africa, the community knowledge about prevention and cause of schistosomiasis had been worryingly low (33, 34). Our study discovered that most participants with the infection had little knowledge about how it is caused, contracted and the health effects associated with it. Therefore, it is cardinal to sensitize the risk community about the disease. Primary school teachers should design deliberate policies to educate learners on the cause, transmission and notable signs/symptoms of schistosomiasis for quicker identification and control of the infection.

The study had its own limitations. Firstly, Maramba stream separates Maramba and Lubuyu compounds but school going children from Libuyu compound were not considered, which could have contributed to the low prevalence rate. Secondly, the stream needed to be assessed for the presence of snail vectors for schistosomes. Thirdly, urinalysis was not conducted especially in infected individuals to assess the extent of urinary tract damage by the disease. Finally, the COVID19 pandemic and the misconception that community had about urine collection led guardians to block urine specimen submission from children. Therefore, more robust studies are necessary to fully comprehend the extent of the disease in all the compounds surrounding Maramba stream.

## Conclusion

The prevalence of urogenital schistosomiasis among school going children in Maramba compound was 3.47% and the correlates of acquiring *S haematobium* included washing in a stream, age and poor knowledge on schistosomiasis. Participants that had schistosomiasis often presented with dysuria and hematuria. Therefore, we recommend that an integrated program that involves sensitizing the community about schistosomiasis and its complications as well as hygiene and behavioral change program could be implemented for easy risk factor modification. The clinic with the help of community health assistants may work hand-in-hand with the community to periodically screening school going children to capture and treat the asymptomatic individuals. Finally, use of molluscicides on Maramba stream to control vector infestation could subsequently control urogenital schistosomiasis.

## Data Availability

None

## Acknowledgements

We would like to graciously acknowledge all parents, guardians and teachers of Maramba school for their unwavering support during data collection, and all the study participants for their cooperation during specimens and data collection. We would like to further acknowledge the contribution of the laboratory technologists and staff members of Maramba clinic for their assistance. Finally, we would like to sincerely thank Dr Benson Hamooya for his advice on data analysis plan. We thank you all for making this study a success.

